# COVID-19: Beliefs in misinformation in the Australian community

**DOI:** 10.1101/2020.08.04.20168583

**Authors:** K Pickles, E Cvejic, B Nickel, T Copp, C Bonner, J Leask, J Ayre, C Batcup, S Cornell, T Dakin, RH Dodd, JMJ Isautier, KJ McCaffery

**Affiliations:** University of Sydney, Sydney Health Literacy Lab, School of Public Health, Faculty of Medicine and Health, NSW 2006 AUSTRALIA; University of Sydney, Susan Wakil School of Nursing and Midwifery, Faculty of Medicine and Health, NSW 2006 AUSTRALIA

**Keywords:** COVID-19, coronavirus, misinformation, myths, digital health literacy, social media

## Abstract

**Objectives:** To investigate prevalence of beliefs in COVID-19 misinformation and examine whether demographic, psychosocial and cognitive factors are associated with these beliefs, and how they change over time.

**Study design:** Prospective national longitudinal community online survey.

**Setting:** Australian general public.

**Participants:** Adults aged over 18 years (n=4362 baseline/Wave 1; n=1882 Wave 2; n=1369 Wave 3).

**Main outcome measure:** COVID-19 misinformation beliefs.

**Results:** Stronger agreement with misinformation beliefs was significantly associated with younger age, male gender, lower education, and primarily speaking a language other than English at home (all p<0.01). After controlling for these variables, misinformation beliefs were significantly associated (p<0.001) with lower digital health literacy, lower perceived threat of COVID-19, lower confidence in government, and lower trust in scientific institutions. The belief that the threat of COVID-19 is “greatly exaggerated” increased between Wave 1-2 (p=0.002), while belief that herd immunity benefits were being covered up decreased (p<0.001). Greatest support from a list of Australian Government identified myths was for those regarding ‘hot temperatures killing the virus’ (22%) and ‘Ibuprofen exacerbates COVID-19’ (13%). Lower institutional trust and greater rejection of official government accounts were associated with greater support for COVID-19 myths after controlling for sociodemographic variables.

**Conclusion:** These findings highlight important gaps in communication effectiveness. Stronger endorsement of misinformation was associated with male gender, younger age, lower education and language other than English spoken at home. Misinformation can undermine public health efforts. Public health authorities must urgently target groups identified in this study when countering misinformation and seek ways to enhance public trust of experts, governments, and institutions.

---

False, misleading or inaccurate health information can pose a serious risk to public health and public action. Misinformation about COVID-19 is common and has spread rapidly across the globe through social media platforms and other information systems.[1–4] In February, the World Health Organisation’s Director-General declared the global ‘over-abundance’ of COVID-19 information an ‘infodemic’.[5] The term ‘misinfodemic’ has since been coined to capture the corresponding rise in misinformation surrounding the virus.

COVID-19 misinformation – which is typically compelling, persuasive, and emotive – spreads significantly farther, faster, deeper, and more broadly than factual information on social media platforms [6]. This is particularly true within tight-knit communities, as has been seen with the spread of vaccine misinformation among some communities in the United States and Sweden. [7–9] Common COVID-19 beliefs circulating in mainstream media include framing of the pandemic as a leaked bioweapon, a consequence of 5G wireless technology, a political hoax, and that the COVID-19 pandemic has been made up by governments to control people. Others detail ineffective measures that individuals can take to prevent or treat the virus, such as exposing themselves to sunlight or taking vitamin C. [10]

Misinformation can undermine public health efforts through shaping beliefs and attitudes - particularly if encountered within a social network - and by reinforcing pre-existing values and positions [11]. Importantly, lower perceived risk or perceived efficacy of prevention behaviours and altered perception of social norms might influence willingness to follow recommendations, such as voluntary testing, isolation and, potentially, vaccination [12].

Understanding what the public know about COVID-19 and identifying beliefs based on misinformation can help shape effective public health communications to ensure efforts to reduce viral transmission are not undermined.

This paper uses data from a longitudinal cohort study of the Australian public to: a) investigate prevalence of beliefs in COVID-19 misinformation in this sample and examine whether any demographic, psychosocial and cognitive factors are associated with these beliefs, and b) investigate how these misinformation beliefs change overtime.

## Methods

### Design

The data used in this study are from a prospective longitudinal national survey exploring variation in understanding, attitudes, and uptake of COVID-19 health advice during the 2020 pandemic lockdown[13, 14]. A total of 4362 participants completed the baseline survey (Wave 1), recruited in Australia in April 2020 (17-24^th^). This survey was administered one month after the first measures of physical distancing and quarantine measures were introduced in Australia and when an increasing number of COVID-19 cases were being recorded. A subset (n=3214) of the sample were invited for longitudinal follow-up to assess changes in attitudes, beliefs and behaviour over the course of the pandemic. Wave 2 of the study (n=1882, 59% response rate) was administered in May (8-15^th^), 3 weeks after the baseline (Wave 1) survey. Wave 3 of the study (n=1369, 43% response rate) was administered in June (5-12^th^), approximately 6 weeks after the baseline survey, when restrictions in Australia showed signs of easing and new case numbers and reported community transmission had fallen dramatically. Wave 3 was administered prior to the resurgence of cases in some areas of Australia.

### Participants and recruitment

Participants were recruited via advertisement on social media (i.e. Facebook, Instagram) and by a market research company (Dynata). Those recruited on social media were invited for longitudinal follow-up. Eligibility criteria included being 18 years or older, currently living in Australia, and able to read and understand English. Participants recruited via Dynata received points for completing the survey which can be redeemed for gift vouchers, donations to charities, or cash. Participants recruited via social media were given the opportunity to enter into a prize draw for the chance to win one of ten $20 gift cards upon completion of each survey wave.

### Ethics approval

Ethics approval was obtained from the Human Research Ethics Committee of The University of Sydney (2020/212). Completion and submission of the online questionnaire was considered evidence of consent.

#### Measures

The survey was built and administered online using Qualtrics (SAP, Provo, UT). Survey items included in each wave were modified from the national longitudinal study [13] to reflect psychological, behavioural, and knowledge factors considered most relevant at that stage of restrictions. Relevant measures for this study are detailed in Table 1. Age, gender, education, language other than English (LOTE) spoken at home, and socioeconomic statues (SES) were assessed in Wave 1 as detailed in [13].

**Table 1.**
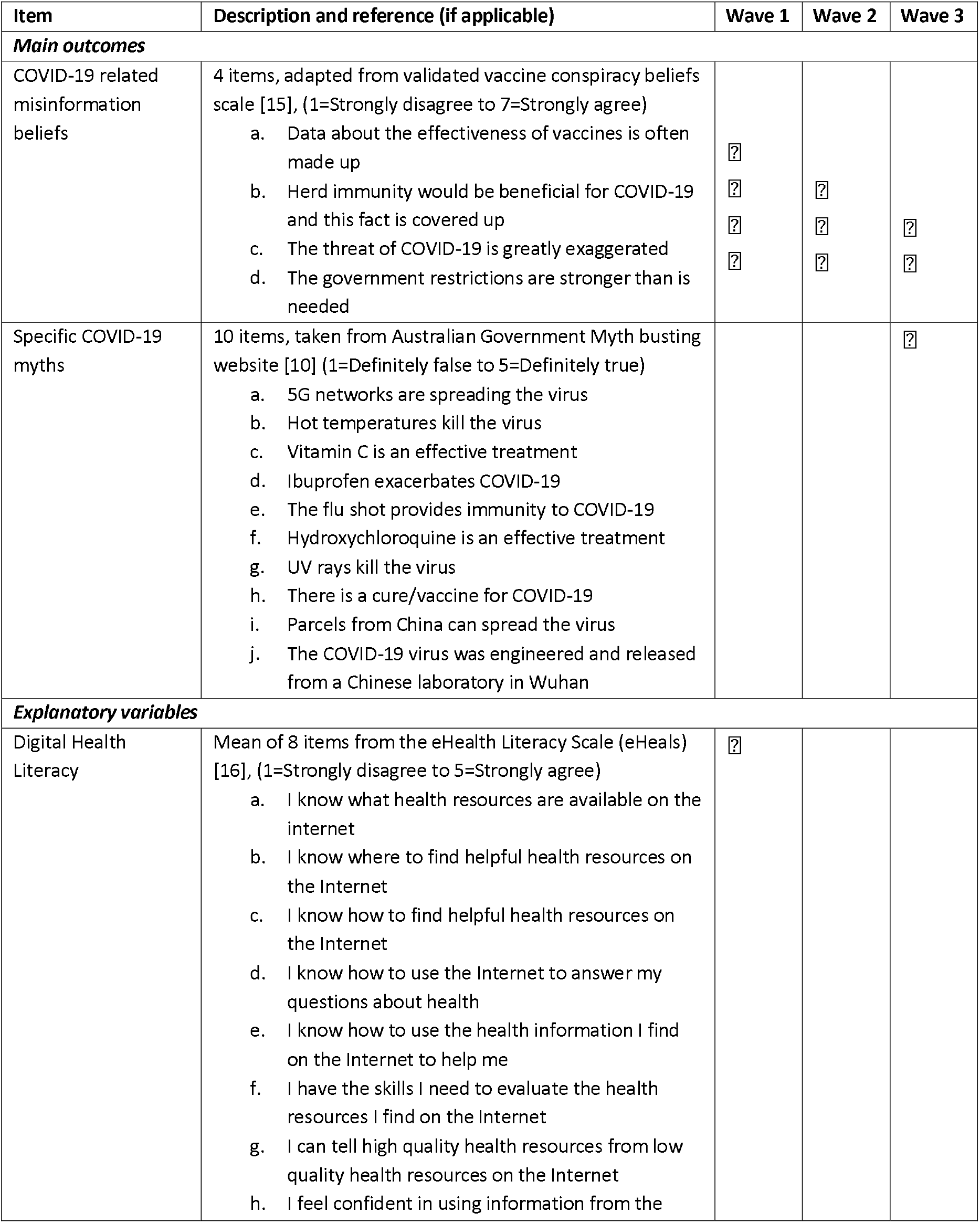
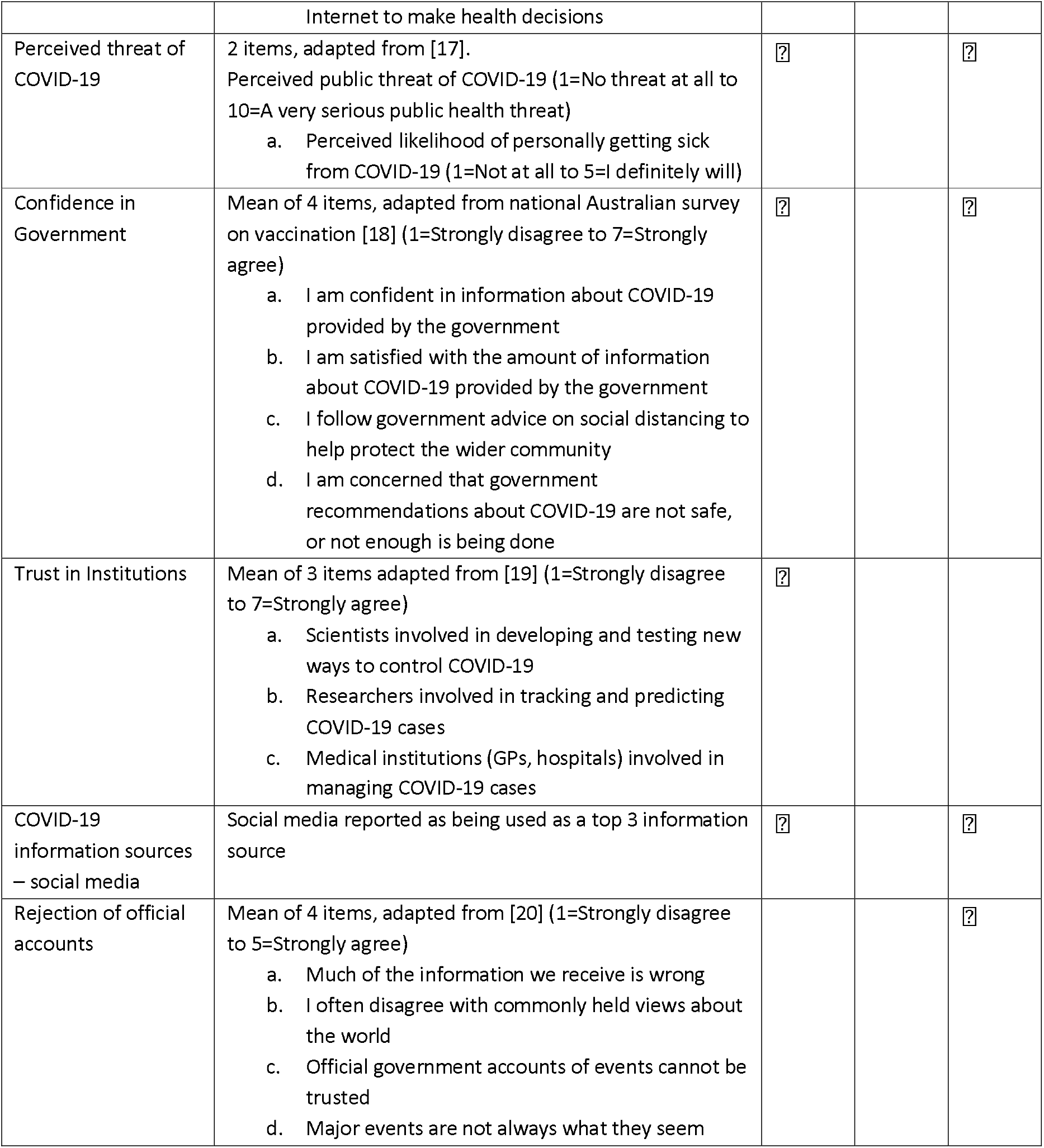
Measures included in this study

### Analysis

Analyses were conducted using Stata/IC v16.1 (StataCorp, College Station, TX). The threshold for statistical significance was set at p<0.05. Descriptive statistics (means and standard deviations for continuous variables, frequency and relative frequency for categorical variables) were calculated for participant characteristics and study outcomes. To reduce the number of outcomes for analysis, misinformation beliefs at baseline were combined into a single measure using principal component analysis (PCA). Associations between the extracted misinformation component and possible explanatory variables were explored using truncated linear regression controlling for sociodemographic factors previously shown to be associated with misinformation beliefs [13].

Changes in misinformation beliefs across study waves were examined using linear mixed models with random intercepts by participant and robust standard errors. Due to changes in the items included in each wave, these items were analysed individually.

Dimension reduction using PCA was applied to the 10 specific COVID-19 myth items (included in Wave 3 of the study). Multivariable truncated regression models were used to examine associations with the extracted components, using the same explanatory variables as for the analysis of misinformation beliefs from Wave 1. Where survey items were repeated in Wave 3 (i.e. perceived threat of COVID-19, confidence in government, and use of social media as a “top 3” information source), this version of the variable was included; otherwise the response at baseline was carried forward (i.e. digital health literacy, institutional trust, and sociodemographic variables). An additional explanatory variable added in Wave 3 (rejection of official accounts) was also included in these models.

## Results

### Sample characteristics

Sample characteristics by each wave are summarised in Table 2.

**Table 2.**
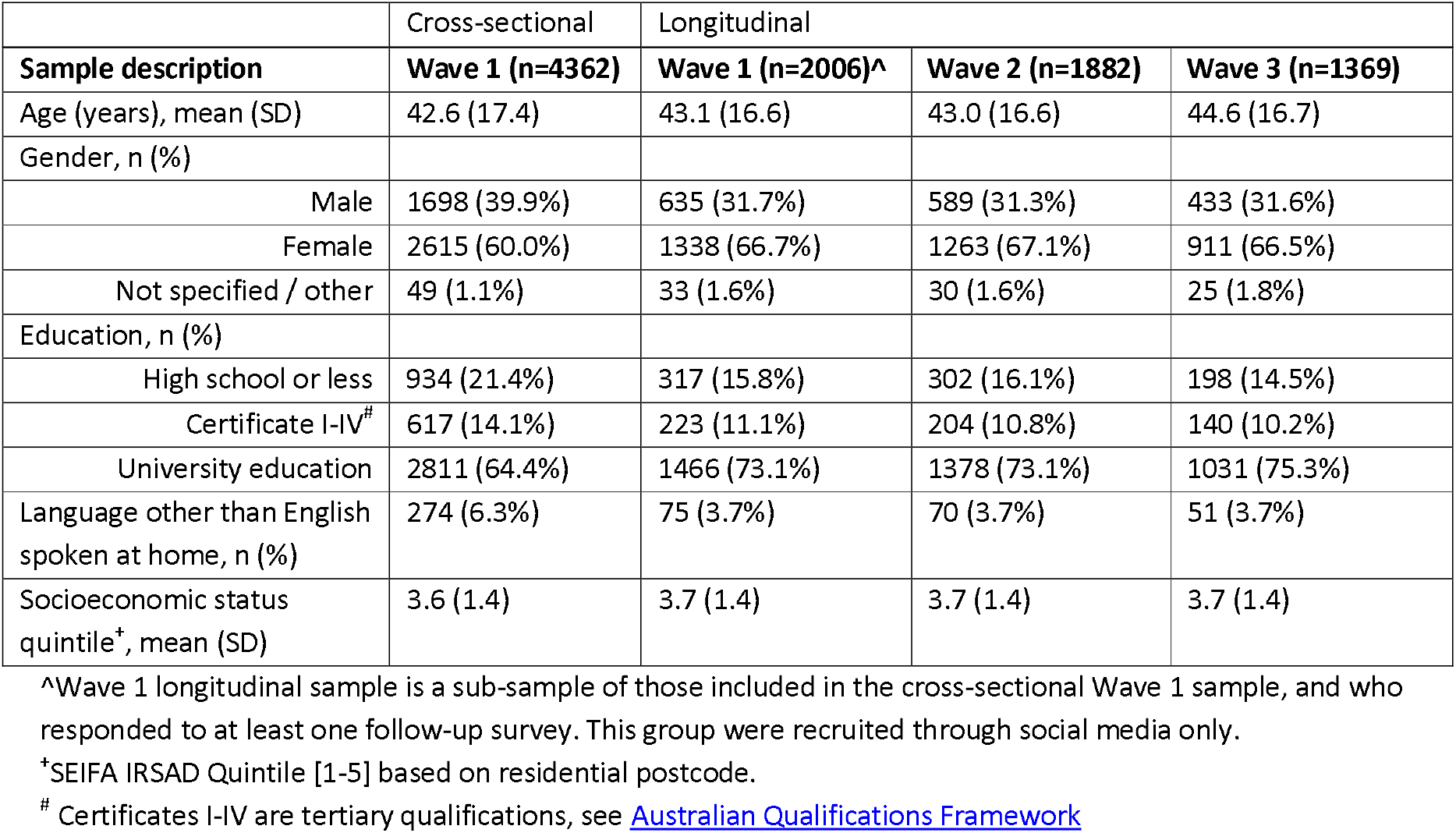
Sample characteristics by study wave (1-3)

|  |  |  |
| --- | --- | --- |
|  | Cross-sectional | Longitudinal |

| Sample description | Wave 1 (n=4362) | Wave 1 (n=2006)^ | Wave 2 (n=1882) | Wave 3 (n=1369) |
| --- | --- | --- | --- | --- |
| Age (years), mean (SD) | 42.6 (17.4) | 43.1 (16.6) | 43.0 (16.6) | 44.6 (16.7) |
| Gender, n (%) |  |  |  |  |
| Male | 1698 (39.9%) | 635 (31.7%) | 589 (31.3%) | 433 (31.6%) |
| Female | 2615 (60.0%) | 1338 (66.7%) | 1263 (67.1%) | 911 (66.5%) |
| Not specified / other | 49 (1.1%) | 33 (1.6%) | 30 (1.6%) | 25 (1.8%) |
| Education, n (%) |  |  |  |  |
| High school or less | 934 (21.4%) | 317 (15.8%) | 302 (16.1%) | 198 (14.5%) |
| Certificate I-IV<sup>#</sup> | 617 (14.1%) | 223 (11.1%) | 204 (10.8%) | 140 (10.2%) |
| University education | 2811 (64.4%) | 1466 (73.1%) | 1378 (73.1%) | 1031 (75.3%) |
| Language other than English spoken at home, n (%) | 274 (6.3%) | 75 (3.7%) | 70 (3.7%) | 51 (3.7%) |
| Socioeconomic status quintile<sup>+</sup>, mean (SD) | 3.6 (1.4) | 3.7 (1.4) | 3.7 (1.4) | 3.7 (1.4) |

<sup>^</sup>Wave 1 longitudinal sample is a sub-sample of those included in the cross-sectional Wave 1 sample, and who responded to at least one follow-up survey. This group were recruited through social media only.
<sup>+</sup>SEIFA IRSAD Quintile [1-5] based on residential postcode.
<sup>#</sup> Certificates I-IV are tertiary qualifications, see Australian Qualifications Framework

### Misinformation beliefs and associations with sociodemographic, cognitive and psychosocial variables (Wave 1)

One month into lockdown in Australia, 753 (17.3%) participants agreed that data about the effectiveness of vaccines is often made up; 652 (15.0%) agreed that that herd immunity would be beneficial for COVID-19 but this is covered up; 603 (13.8%) agreed that the threat of COVID-19 is greatly exaggerated; and 595 (13.6%) agreed that the Australian government restrictions are stronger than required. Responses on these items were moderately correlated (pairwise r’s between 0.36 and 0.63), with good internal consistency (Cronbach’s α= 0.78) and sufficient sampling adequacy (KMO = 0.76). PCA of these items resulted in the extraction of a single component accounting for 60.7% of the variance (component loadings provided in Appendix Table 1).

Estimated marginal means from the multivariable regression model of misinformation beliefs at baseline are provided in Table 3. Stronger agreement with misinformation beliefs were significantly associated with younger age, male gender, lower education, and primarily speaking a language other than English at home (all p<0.01). After controlling for these variables, misinformation beliefs were significantly associated (p<0.001) with lower digital health literacy, lower perceived threat of COVID-19, lower confidence in government, and lower trust in scientific institutions.

**Table 3:** Multivariable truncated linear regression of strength of agreement with misinformation beliefs. Higher values of the outcome indicate greater support for misinformation. Data are presented as estimated marginal mean differences (95% CIs).

| Explanatory variables |  | Estimated marginal mean differences (95% confidence interval) |
| --- | --- | --- |
| **Sociodemographic variables** |  |  |
| Age (/year) |  | **-0.023 (-0.028, -0.018)\*\*\*** |
| Female gender (vs male) |  | **-0.384 (-0.541, -0.226)\*\*\*** |
| Education (vs high school or less) |  |  |
| Certificate I-IV<sup>#</sup> |  | 0.114 (-0.133, 0.360) |
| University education |  | **-0.270 (-0.459, -0.080)\*\*** |
| LOTE |  | **0.847 (0.569, 1.126)\*\*\*** |
| SES (/quintile) |  | -0.050 (-0.105, 0.005) |
| **Additional explanatory variables** | **Sample means** |  |
| Digital health Literacy (1-5) | 4.04 (0.74) | **-0.250 (-0.356, -0.144)\*\*\*** |
| Perceived public threat of COVID-19 (1-10) | 7.64 (2.17) | **-0.336 (-0.372, -0.300)\*\*\*** |
| Not likely to get sick (n, %) | 1091 (25.5%) | **0.649 (0.475, 0.823)\*\*\*** |
| Mean confidence in government (1-7) | 5.15 (1.06) | **-0.143 (-0.222, -0.063)\*\*\*** |
| Mean institutional trust (1-7) | 5.95 (1.06) | **-0.663 (-0.738, -0.587)\*\*\*** |
| Social media used as a top 3 information source (n, %) | 1923 (44.9%) | 0.151 (-0.001, 0.307) |

<sup>+</sup> marginal mean differences are not reported for gender = not specified or other due to small sample size but was included in the regression model. Statistical significance is indicated by \* $p < 0.05$ ; \*\* $p < 0.01$ ; \*\*\* $p < 0.001$ . $n = 4286$ complete cases included in analyses; missing values were not imputed.
<sup>#</sup> Certificates I-IV are tertiary qualifications, see Australian Qualifications Framework

### Changes in misinformation beliefs over time

Prevalence of agreement with misinformation by study wave is shown in Table 4, which appear to be generally consistent over time. Linear mixed models identified a main effect of time (p=0.006), with pairwise contrasts showing an increase in the belief that the threat of COVID-19 is greatly exaggerated between Wave 1 and Wave 2 (Item 3; mean difference [MD] = 0.079, 95%CI: 0.030, 0.128, p=0.002). There was a decrease in the belief that herd immunity is beneficial for COVID-19, but is covered up, between Wave 1 and Wave 2 (Item 2; MD = −0.130, 95%CI: −0.189, −0.071, p<0.001). There was no difference across time for Item 4 (strength of government restrictions; p=0. 41).

**Table 4.** Estimated means (95% confidence intervals) and prevalence of agreement (i.e., responded as somewhat agree [5] to strongly agree [7]) with misinformation beliefs by study wave.

|  | **Wave 1 (baseline)**<br>**(n=2006)** |  | **Wave 2**<br>**(n=1882)** |  | **Wave 3**<br>**(n=1369)** |  |
| --- | --- | --- | --- | --- | --- | --- |
| Items<br>[scale range 1-7] | Mean<br>(95% CI) | Agree<br>(n; %) | Mean<br>(95% CI) | Agree<br>(n; %) | Mean<br>(95% CI) | Agree<br>(n; %) |
| Data about the effectiveness of vaccines is often made up | 2.37<br>(2.30, 2.44) | 230 (11.5%) | Not assessed |  |  |  |
| Herd immunity would be beneficial for COVID-19 and this fact is covered up | 2.52<br>(2.46, 2.59) | **213 (10.6%)** | 2.39<br>(2.32, 2.46) | **185 (9.8%)** | Not assessed |  |
| The threat of COVID-19 is greatly exaggerated | 1.99<br>(1.93, 2.05) | **182 (9.1%)** | 2.07<br>(2.00, 2.14) | **185 (9.8%)** | 2.04<br>(1.98, 2.10) | 110 (8.0%) |
| Government restrictions are stronger than is needed | 2.14<br>(2.08, 2.21) | 197 (9.8%) | 2.16<br>(2.09, 2.22) | 186 (9.9%) | 2.19<br>(2.12, 2.25) | 125 (9.1%) |

### Specific misinformation beliefs and associations with sociodemographic, cognitive and psychosocial variables (Wave 3 only)

The level of agreement across the 10 COVID-19 myths from the Australian Government website had moderate internal consistency (Cronbach’s α= 0.693), and sufficient sampling adequacy (KMO = 0.761). Application of PCA (with varimax rotation) identified a three-component solution which cumulatively accounted for 51.15% of the variance (see Appendix Table 2 for component loading and proportion agreeing with each item); examination of the contributing items to each component resulted in the following 3 labels:

1. Symptom management and prevention myths (PC1; explaining 18.9% of the total variance)
2. Causes and transmission myths (PC2; explaining 16.7% of the total variance); and
3. Immunity and cure myths (PC3; explaining 15.6% of the total variance).

Regarding specific myths concerning *symptom management and prevention*, 301 (22%) of the Wave 3 sample (n=1369) agreed that hot temperatures kill the virus, 295 (21.5%) agreed that UV rays kill the virus, and 179 (13.1%) agreed that Ibuprofen exacerbates COVID-19 (see Appendix Table 2). Greater support for symptom management and prevention myths (PC1) was significantly associated with younger age and male gender; and after controlling for demographics (age, gender, education and LOTE) was associated with lower institutional trust and greater rejection of official accounts (PC1; see Table 5).

For myths regarding *causes and transmission*, 167 (12.2%) of the sample agreed the COVID-19 virus was engineered and released from a Chinese laboratory in Wuhan, 57 (4.2%) agreed that parcels from China can spread the virus, and only 8 (0.6%) agreed that 5G networks are spreading the virus. Causes and transmission myths (PC2) were significantly associated with less education and more social disadvantage. Greater belief in these myths was associated with lower digital health literacy, reduced perceived public threat, reduced institutional trust, and greater rejection of official accounts after controlling for sociodemographic variables (PC2; see Table 5).

Regarding myths about *immunity and cure*, 62 (4.5%) of the sample agreed Vitamin C is an effective treatment, 55 (4.0%) agreed there is a cure/vaccine for COVID-19, 32 (2.3%) agreed Hydroxychloroquine is an effective treatment, and 15 (1.1%) agreed the flu shot provides immunity. Greater support for immunity and cure myths (PC3) was significantly associated with younger age. After controlling for sociodemographic factors, lower digital health literacy, reduced perceived public threat, reduced institutional trust, and greater rejection of official accounts were associated with greater belief in these myths (PC3; see Table 5).

**Table 5:** Multivariable truncated linear regression of the misinformation beliefs (n=1366). Higher values of the outcome indicate greater support for these beliefs. Data are presented as estimated marginal mean differences (95% confidence intervals)

|  |  | Symptom management & prevention (PC1) | Causes & transmission (PC2) | Immunity & cure (PC3) |
| --- | --- | --- | --- | --- |
| **Sociodemographic variables<sup>^</sup>** |  | Estimated marginal mean difference (95% confidence interval) |  |  |
| Age (/year) |  | **-0.007 (-0.014, -0.001)\*** | 0.005 (-0.003, 0.014) | **-0.021 (-0.029, -0.013)\*\*\*** |
| Female gender (vs male)<sup>+</sup> |  | **-0.397 (-0.610, -0.184)\*\*\*** | 0.222 (-0.087, 0.530) | -0.088 (-0.341, 0.165) |
| Education (vs high school or less) |  |  |  |  |
| Certificate I-IV<sup>#</sup> |  | 0.155 (-0.245, 0.556) | 0.401 (-0.109, 0.912) | 0.266 (-0.185, 0.716) |
| University education |  | 0.173 (-0.121, 0.467) | **-0.498 (-0.899, -0.096)\*** | -0.051 (-0.388, 0.285) |
| LOTE |  | -0.298 (-0.827, 0.230) | 0.463 (-0.254, 1.18) | 0.212 (-0.375, 0.799) |
| SES (/quintile) |  | -0.032 (-0.104, 0.040) | **-0.212 (-0.313, -0.111)\*\*\*** | -0.007 (-0.092, 0.079) |
| **Additional explanatory variables** | **Sample means** |  |  |  |
| Digital health Literacy (1-5)<sup>^</sup> | 4.18 (0.67) | 0.105 (-0.046, 0.255) | **-0.304 (-0.512, -0.097)\*\*** | **-0.444 (-0.618, -0.270)\*\*\*** |
| Perceived public threat of COVID-19 (1-10) | 7.33 (2.44) | -0.027 (-0.069, 0.016) | **-0.074 (-0.133, -0.015)\*** | **-0.057 (-0.107, -0.007)\*** |
| Not likely to get sick (n, %) | 123 (9.0%) | 0.083 (-0.264, 0.429) | -0.285 (-0.781, 0.211) | 0.133 (-0.267, 0.535) |
| Mean confidence in government (1-7) | 5.52 (0.94) | 0.028 (-0.094, 0.149) | 0.117 (-0.054, 0.288) | 0.051 (-0.093, 0.194) |
| Mean institutional trust (1-7)<sup>^</sup> | 6.15 (0.95) | **-0.229 (-0.339, -0.119)\*\*\*** | **-0.599 (-0.750, -0.448)\*\*\*** | **-0.226 (-0.353, -0.099)\*\*\*** |
| Social media used as a top 3 information source (n, %) | 680 (49.8%) | 0.107 (-0.094, 0.307) | 0.200 (-0.087, 0.486) | 0.177 (-0.063, 0.417) |
| Rejection of official accounts (1-5) | 2.36 (0.83) | **0.172 (0.031, 0.313)\*** | **0.451 (0.245, 0.657)\*\*\*** | **0.337 (0.169, 0.506)\*\*\*** |

<sup>^</sup> indicates values that were obtained at baseline (wave 1) and carried forward
<sup>+</sup> marginal mean differences are not reported for gender = not specified or other due to small sample size but were included in the regression model. Statistical significance is indicated by \*p<0.05; \*\* p<0.01; \*\*\*p<0.001. Analysis sample n=1366; missing data were not imputed.
<sup>#</sup> Certificates I-IV are tertiary qualifications, see Australian Qualifications Framework

## Discussion

Our findings showed lower institutional trust, lower digital health literacy, and greater rejection of official accounts were associated with stronger support of misinformation beliefs. Misbeliefs were also more common among those who primarily spoke a language other than English at home, in younger age groups, and in males. The most commonly held misinformation beliefs in this study concerned symptom management and prevention. We found small changes between wave 1 and wave 2 on two of the beliefs: an increase in the belief that COVID-19 is greatly exaggerated and a decrease in the belief that herd immunity is beneficial for COVID-19 but is covered up. Despite these differences being statistically significant, they likely have little to no practical importance (i.e., only a 0.08 and 0.12 unit change respectively on a 7-point scale). Notably, the proportion of participants agreeing with each item remained generally consistent overtime.

The study was large and diverse but not representative of the national population. Given this, caution is needed in generalising from the prevalence findings. However, we have identified factors associated with misinformation beliefs and trends over time for some variables.

Our rates of COVID-19 misinformation beliefs were lower when compared internationally [21] [22], An Australian poll conducted in May 2020 found relatively high support (12-77%) for misinformation beliefs relating to the creation, spread, and prevention of the virus [23]. Interestingly, we saw a much lower prevalence of people agreeing that 5G networks are spreading the virus when compared with the Essential Research findings.

The poll found similar demographic patterns to our findings, where men and younger people agreed with a range of COVID-19 myths more than other groups. In the US and UK, younger people are more likely to hold conspiracy beliefs about COVID-19 [24] [25]. American men are more likely to agree with COVID-19 conspiracy theories than women [26].

The association between misinformation beliefs and lower education, language other than English, younger age and male gender point to important gaps in public health messaging to these groups. We have previously highlighted these disparities [14] and the complexity of government health information about COVID-19. In our previous analysis we found that some of these groups (people with less education, language other than English) had poorer understanding of COVID-19 symptoms and were less able to identify behaviours to prevent infection. Recent attention has focused on the importance of reaching people who do not speak English as a first language [27], Our study further highlights the need for health information to be written to meet diverse health literacy requirements and targeted to specific groups. Young people should be involved in the design of COVID-19 messages to get the tone and delivery right. This can be done through end user testing, consumer focus groups, and including young people on communication teams.

Provision of quality information online is unlikely to be a sufficient strategy to counter the influence of misinformation if digital health literacy is not accounted for. Messaging and debunking must be delivered on multiple channels, consistent in content and style, and conveyed in local languages to ensure engagement with all communities. [28] Emerging evidence supports the idea that prebunking or ‘innoculating’ people against misinformation can build resistance to misinformation across cultures. [29] Going forward, it will be important to invest in programs teaching digital health literacy and healthy scepticism of health news. Finally, partnerships between public health authorities and trusted organisations to deliver information and correct misinformation should be utilised where possible. [30] Corrective messages are most successful when they offer a coherent explanation for how and why a belief based on misinformation is incorrect.[31] Research shows that corrective information can counter misperceptions and improve belief accuracy after a person’s exposure to misinformation.[32]

Timely, accurate and transparent messaging is vitally important to gaining public trust in messaging from authorities ahead of other less credible sources [33]. While there is now intense global interest aimed at limiting the spread of misinformation in the first place,[1, 29, 34] it will require ‘a sustained and coordinated effort by independent fact checkers, independent news media, platform companies, trusted spokespeople and public authorities to help the public understand and navigate the pandemic’.[35] Government health agencies and news media are ideally positioned to successfully correct misbeliefs during public health crises.[32]

Around the world and in Australia, anti-lockdown protests have taken place in capital cities, with protesters voicing opposition to vaccination, telecommunication towers, and the COVID ‘hoax’. Researchers have recently investigated the degree to which misinformation about COVID-19 is associated with people’s willingness to adhere to public health recommendations and government enforced measures and found that willingness decreases significantly as the strength of misbeliefs increases,[36, 37] including decreased COVID-19 vaccination intentions [38]. In some cases, misinformation has led to serious harm, such as the Iranian methanol poisoning experience [39]. The spread of misinformation is an ongoing area of concern as Australia and the world continue to live with the fluctuating realities of a global pandemic. Correcting misinformation should be viewed as a vitally important science and health policy activity.[40]

### Strengths & limitations

This was a large and diverse sample of the Australian population and the longitudinal design enabled us to look at whether misinformation beliefs changed over the course of the pandemic. By design, the survey items changed across time, however this prevented us from being able to calculate longitudinal changes in the principal component derived at baseline. The sample was recruited via an online panel and social media, the majority were well educated, with a low proportion of culturally and linguistically diverse participants. A final limitation is the labelling of the misinformation items as these particular beliefs are likely contextual and subjective.

## Conclusion

These findings highlight important gaps in communication effectiveness. Misinformation can undermine public health efforts. Public health authorities must urgently target groups identified in this study when countering misinformation and seek ways to enhance public trust of experts, governments, and institutions. Communicators must pay attention to pitching COVID-19 messages at a level that most people can understand, and suitable and relevant to diverse cultural groups, young people and particularly targeted to men.

## Data Availability

The data that support the findings of this study are available from the corresponding author, KP, upon reasonable request.

## Acknowledgements

The SHLL group thanks the participants of the longitudinal COVID-19 survey for their ongoing participation in this research.

## Appendix

**Table 1:** Misinformation beliefs at baseline (wave 1) PC loading

| Item | Component Loading |
| --- | --- |
| Data about the effectiveness of vaccines is often made up | 0.429 |
| The threat of COVID-19 is greatly exaggerated | 0.546 |
| Herd immunity would be beneficial for COVID-19 and this fact is covered up | 0.497 |
| Australian government restrictions are stronger than is needed | 0.521 |

**Table 2.** Agreement^^^ and summary statistics for COVID-19 myth items (selected from the Australian Government COVID-19 Mythbusting website) at Wave 3 (n=1369); and descriptive statistics and component loadings^%^ from Principal Component Analysis.]

| Items [1 to 5] | Disagree, n (%) | Unsure, n (%) | Agree, n (%) | Mean (SD) | Component loadings |  |  |
| --- | --- | --- | --- | --- | --- | --- | --- |
|  |  |  |  |  | PC1 | PC2 | PC3 |
| 5G networks are spreading the virus | 1328 (96.9%) | 356 (2.5%) | 8 (0.6%) | 1.10 (0.43) |  | 0.590 |  |
| Hot temperatures kill the virus | 808 (58.9%) | 262 (19.1%) | 301 (22.0%) | 2.30 (1.28) | 0.596 |  |  |
| Vitamin C is an effective treatment | 1128 (82.3%) | 181 (13.2%) | 62 (4.5%) | 1.62 (0.90) |  |  |  |
| Ibuprofen exacerbates COVID-19 | 746 (54.4%) | 446 (32.5%) | 179 (13.1%) | 2.33 (1.07) | 0.351 |  |  |
| The flu shot provides immunity to COVID-19 | 1317 (96.1%) | 39 (2.8%) | 15 (1.1%) | 1.17 (0.51) |  |  | 0.658 |
| Hydroxychloroquine is an effective treatment | 1008 (73.5%) | 331 (24.1%) | 32 (2.3%) | 1.85 (0.87) |  |  |  |
| UV rays kill the virus | 734 (53.5%) | 342 (25.0%) | 295 (21.5%) | 2.39 (1.25) | 0.618 |  |  |
| There is a cure/vaccine for COVID-19 | 1218 (88.8%) | 98 (7.2%) | 55 (4.0%) | 1.37 (0.81) |  |  | 0.629 |
| Parcels from China can spread the virus | 1179 (86.0%) | 135 (9.8%) | 57 (4.2%) | 1.66 (0.82) |  | 0.460 |  |
| The COVID-19 virus was engineered and released from a Chinese laboratory in Wuhan | 1008 (73.5%) | 196 (14.3%) | 167 (12.2%) | 1.95 (1.09) |  | 0.620 |  |

<sup>^</sup> Items were recoded to disagree [1-2], neutral/unknown [3], agree [4-5]. <sup>%</sup> Component loadings less than |0.3| have been omitted.

